# Reverse causal effect of atrial fibrillation on 17 site-specific cancer risk: A Mendelian randomization study

**DOI:** 10.1101/2021.01.10.21249534

**Authors:** Sehoon Park, Soojin Lee, Yaerim Kim, Semin Cho, Kwangsoo Kim, Yong Chul Kim, Seung Seok Han, Hajeong Lee, Jung Pyo Lee, Kwon Wook Joo, Chun Soo Lim, Yon Su Kim, Dong Ki Kim

## Abstract

**Background:** Bidirectional association between atrial fibrillation (AF) and cancer was reported by observational investigations. Additional study is warranted to investigate the causal effects of AF on risk of cancer.

**Methods:** This study was a summary-level Mendelian randomization (MR) analysis. Genetic instrument for AF was developed from a genome-wide association study (GWAS) meta-analysis for AF including 537,409 European ancestry individuals including 55,144 AF cases. The outcome data for risk of 17 site-specific cancer from a previous GWAS meta-analysis of the UK Biobank (48,961/359,825 case/controls) and Genetic Epidemiology Research on Adult Health and Aging (16,001/50,525 case/controls) cohorts including European ancestry individuals was investigated. Inverse variance weighted method was the main MR method, supported by pleiotropy-robust sensitivity analysis including MR-Egger regression and penalized weighted median method.

**Results:** The causal estimates indicated that AF was causally linked to higher risk of cancers of lung, breast, cervix, endometrium, and melanoma. MR-Egger test for directional pleiotropy indicated absence of a pleiotropy in the identified causal estimates and MR-Egger regression and median-based methods provided similarly significant findings. On the other hand, the genetic predisposition of AF was significantly associated with lower risk of esophagus/gastric cancer, but possibility of a directional pleiotropy remained in the association.

**Conclusions:** AF is a causal factor for certain types of cancer. Appropriate cancer screening should be suggested in clinical guidelines for AF patients. Future trial is necessary to confirm whether appropriate management of AF may reduce the risk of cancer which is a major cause of deaths in AF patients.

## Introduction

Atrial fibrillation (AF) is the most prevalent cardiac arrhythmia worldwide.^1^ Socioeconomic burden related to AF is substantial and further project to increase in the future along with global ageing trends. AF is a direct etiology for thromboembolism, causes compromise in cardiac output, and is associated with various health outcomes including cardiovascular diseases and mortality.

Cancer, another leading causes of mortality,^2^ is commonly prevalent with AF in elderly individual and is a common cause of death in AF patients. Previous reports suggested that cancer increases the risk of AF through certain mechanisms.^3^ However, a reverse-association has also been suggested, showing that presence of AF is an independent risk factor for future cancer.^4-6^ However, as the findings are based on observational studies and delayed diagnosis of previous subclinical cancer is possible, the issue of reverse causation hinders the causal interpretation of the associations of AF and risk of cancer. Furthermore, as AF and cancer are both prevalent in elderly individuals commonly with multiple comorbidities, unmeasured confounding effect may bias the previous observational findings. Therefore, additional study is warranted to investigate the causal effects of AF on site-specific cancer risk, which would aid establishing new guidelines for cancer screenings in the emerging population with AF.^4^

Mendelian randomization (MR) is an analytic method to demonstrate causal effect by implementing genetic instruments.^7^ As genetic instrument is determined before birth, causal estimates from genetic predisposition to an outcome are minimally affected by reverse causation or confounding effects. As those with a higher genetic predisposition for an exposure of interest would actually have higher prevalence of the exposure, a significant association between the genetic predisposition and outcome suggests a presence of a causal effect. MR has been widely adopted in recent medical literature and identified important causal pathways between various exposures and complex health outcomes.^8-10^ In this study, we aimed to investigate the causal effect of AF on 17 site-specific cancer risks by MR analysis. We implemented large-scale genome-wide association study (GWAS) results for AF and various types of cancers and performed a large-scale summary-level MR analysis.

## Methods

### Ethical considerations

The study was approved by the Institutional Review Boards of Seoul National University Hospital (IRB No. E-2012-004-1177). Our investigation for the UK Biobank data base has been approved by the consortium (application No. 53799). The requirement for informed consent was waived because the study analyzed public GWAS results and anonymous databases.

### Study setting

The study was a summary-level MR. To limit the effects from different ethnic distributions in a genetic analysis, the studied data were restricted to those from European ancestry. The genetic instrument was developed from the previous GWAS meta-analysis for AF.^11^ The individual-level data of the UK Biobank was implemented to identify genetic variants associated with potential confounders.^12^ The outcome summary statistics for 17 site-specific cancer risks were provided from a previous GWAS meta-analysis, including UK Biobank and Genetic Epidemiology Research on Adult Health and Aging (GERA) cohorts (URL: https://github.com/Wittelab/pancancer_pleiotropy).^13^

### Genetic instrument

The previous GWAS meta-analysis identified single nucleotide polymorphisms (SNPs) showing genome-wide significant association with paroxysmal/persistent AF (Table 1).^11^ The study meta-analyzed GWAS results from AFGen consortium Broad AF study, UK Biobank, and Biobank Japan. The SNPs reported in the study was relevant as being enriched in cardiac structural and electrophysiological pathways. Along with the findings from multi-ethnic population, the study reported 84 independent sentinel SNPs from the European ancestry individuals, including 537,409 European ancestry individuals including 55,144 AF cases. We constructed the genetic instrument from the 84 SNPs and Steiger filtering ensured that the genetic effects are from AF to outcome.^14^ During trimming of the instrument, we considered the following core assumptions of MR.^7^

**Table 1.** Summary of the study population of the previous genome-wide association study meta-analysis.

| Component of this study | Phenotype | Included cohort | Study population which derived the summary statistics | Remarks |
| --- | --- | --- | --- | --- |
| Genetic instrument | Paroxysmal/persistent AF | Summary statistics from AFGen consortium Broad AF study, UK Biobank, and Biobank Japan | 537,409 European ancestry individuals with 55,144 AF cases | Identified independent 84 sentinel SNPs with a genome-wide significant level $P < 1 \times 10^{-8}$) association with AF risk |
| Outcome summary statistics<sup>a</sup> | First identified site-specific cancer | UK Biobank | 408,786 European ancestry individuals with 48,961 cancer cases | Cancer types were grouped to 17 site-specific cancer types to secure statistical power. Testis cancer data was obtained only in the UK Biobank due to data availability. |
|  |  | Genetic Epidemiology Research on Adult Health and Aging (GERA) | 66,526 European ancestry individuals with 16,001 cancer cases |  |

AF = atrial fibrillation, SNPs = single nucleotide polymorphisms
<sup>a</sup>The patient age and cancer grade/stage was various at the time of cancer diagnosis in the cohorts which derived the outcome summary statistics. The detailed information according to the types of site-specific cancer is available in the supplemental material attached to the previously published study by Sara R. Rashkin et al. (URL: https://static-content.springer.com/esm/art%3A10.1038%2Fs41467-020-18246-6/MediaObjects/41467_2020_18246_MOESM1_ESM.pdf).

First, the relevance assumption means that the genetic instrument should be strongly associated with the exposure of interest. The previous GWAS meta-analysis already provided independent SNPs strongly associated with AF and we confirmed the association within the individual-level UK Biobank data by validating the explained variance by allele scores calculated from the genetic instrument.

Second, the independence assumption means that the genetic instrument should not be associated with confounders. We disregarded the SNPs that showed strong (P < 1×10^−5^) association with phenotypical hypertension, diabetes mellitus, obesity, dyslipidemia, thyroid disorder,^10^ and smoking by performing GWAS, with PLINK 2.0, with the individual-level UK Biobank data.^15^ In the GWAS, the linear regression model was constructed adjusted for age, sex, age×sex, age^2^, and the first 10 principal components. Furthermore, we performed additional sensitivity MR analyses which are robust to pleiotropic effects and relax this second assumption for the genetic instruments.^16,17^

Third, the exclusion-restriction assumption means that the genetic effect should be through the exposure of interest. There is yet a formal test for this assumption, but median-based MR method, which was performed as a sensitivity MR analysis, can relax this assumption in up to half of the genetic instruments.^17^

### Individual-level UK Biobank data

UK Biobank is a population-scale data including genetic and various phenotypical information. The UK Biobank included > 500,000 individuals aged 40-69 years across the United Kingdom, and the details have been published previously.^12,18^ In this analysis, we included unrelated 337,138 white British UK Biobank participants after excluding those who were outliers due to heterozygosity or missing rates or had sex chromosome aneuploidy as the individual-level data.^8^ The data has been utilized to identify confounder associated SNPs and to confirm whether the genetic instrument successfully predicted phenotypical AF. In the data, history of diabetes mellitus was self-reported, and hypertension or dyslipidemia history was identified through usage of relevant medication. Obesity was determined by baseline body mass index ≥ 30 kg/m^2^. Thyroid disease was identified by relevant diagnostic codes. The phenotypical AF was determined from the hospital records or death certificates including main causes of deaths, identified by an ICD-10 diagnostic code of I48 or ICD-9 diagnostic code of 4273.

### Outcome GWAS summary statistics for site-specific cancer

The previous GWAS meta-analysis for pan-cancer risk was performed with European ancestry individuals in the UK Biobank and GERA cohort (Table 1).^12,13,19^ The study investigated 48,961/359,825 case/controls in the UK Biobank data and 16,001/50,525 case/controls in the GERA data. The study provided summary statistics for cancers of oropharynx, thyroid, lung, breast, pancreas, esophagus/stomach, colon, rectum, prostate, kidney, bladder, cervix, endometrium, ovary, lymphocytic leukemia, melanoma, and non-Hodgkin lymphoma. In the UK Biobank data, cancer histories were identified through national cancer registries in the United Kingdom collecting cancer cases with medical records, death certificates, and other sources from early 1970 to 2015. In the GERA data, the participants were receiving Kaiser Permanente Nothern (KPNC) health care plan and the registry collected treatment/diagnosis history of cancer from 1988 to 2016. In the meta-analysis, the first cancer diagnosis determined the cancer case, and the individuals without any cancer history constructed the control group. Those with a cancer history which was not the type of interest in the meta-analysis were excluded. Along with novel SNPs predicting site-specific cancer risks, the study had strength that previously reported SNPs in the literature were replicated in the results.

### MR methods

We performed summary-level MR analysis. During harmonization of the data, overlapping SNPs between the genetic instrument and the outcome data were utilized, and we conservatively excluded the palindromic SNPs because whether the effect sizes were from the alleles in the same DNA strand may be questioned for such variants. We performed multiplicative random effect inverse variance weighted method as the main MR method, which allows balanced pleiotropic effect when heterogeneity among the instrument is present.^20^ Next, we performed MR-Egger regression analysis with bootstrapped standard error to yield pleiotropy-robust causal estimates.^16^ The MR-Egger regression allows pleiotropic effect for all of the genetic instruments but still can calculate valid causal estimates. In addition, MR-Egger intercept reflects the presence of directional pleiotropy, thus can be implemented as a formal test for a pleiotropic effect. The weakness of the MR-Egger regression method is the weak statistical power and the method can be biased when group of instruments act through a same pleiotropic pathway [violation of the nstrument Strength Independent of Direct Effect (InSIDE) assumption],^21^ thus additional median-based sensitivity analysis is encouraged to validate the results. Therefore, we performed penalized weighted median method,^17^ which provides valid causal estimates even up to half of the genetic instruments are invalid. The method has particular strength as it can relax the untestable exclusion-restriction assumption in 50% of the genetic instruments and less biased by violation of the InSIDE assumption. Then, we performed MR-PRESSO analysis, which detects and corrects the effects from outliers.^22^ The causal estimates were calculated by MR-PRESSO when the analysis identified a correctable outlier effect. Within instrument heterogeneity was identified through Cochran’s Q statistics P value.

Lastly, additional sensitivity analysis was performed considering the sample overlap (UK Biobank data) in the GWAS for genetic instrument develop and for for derivation of outcome summary statistics.^23^ Such overlap may cause bias towards confounding effects, particularly when the instrumental power is weak. Following the previous literature,^23^ we performed sensitivity analysis by including fewer SNPs but with stronger association strength, by including SNPs with P < 1×10^−10^ association with AF as the genetic instrument. A significant causal effect was addressed only when the inverse variance weighted method and all sensitivity MR methods provided consistently significant (two-sided P value < 0.05) causal estimates. The summary-level MR was performed by the TwoSampleMR package in R (version 3.6.2, the R foundation).^24^

## Results

### Genetic instrument construction

Of the 84 SNPs, there were 4 SNPs showing strong association with the potential confounders. After additionally disregarding non-overlapping or palindromic SNPs, 67 SNPs constructed the genetic instrument for AF. The explained variance for phenotypical AF by the allele scores for AF in the individual-level UK Biobank data was 2.4% (McFadden’s pseudo-R square).

### Summary-level MR results

The summary-level MR results are graphically shown in Figure 2 and the numbers are present in Table 2. A genetically predicted AF was significantly associated with higher risk of cancers in lung, breast, cervix, endometrium, and melanoma, supported by all sensitivity analysis results. In the causal estimates for the cancers significantly affected from genetic predisposition of AF, MR-Egger test for directional pleiotropy or Cochran’s Q statistics did not identify a significant pleiotropic effect or a heterogeneity. Even when we reduced the number of SNPs as the genetic instruments by including only 47 SNPs with P < 1×10^−10^ association with AF, the findings were consistent both by the inverse variance weighted method and the pleiotropy-robust MR analyses (Supplemental Table 1). The MR-PRESSO analysis did not detect correctable outlier effects, thus, calculating the causal estimates from MR-PRESSO was unnecessary.

**Table 2.** Summary-level MR results

| Site-specific cancer | Number of cases (UK Biobank /GERA) | Cochran's Q statistics P value | MR-Egger test P value for directional | MR method | OR (95% CI) | P |
| --- | --- | --- | --- | --- | --- | --- |
| Oropharynx | 930/293 | 0.545 | 0.069 | Inverse variance weighted | 0.996 (0.984, 1.007) | 0.475 |
|  |  |  |  | MR-Egger | 1.003 (0.988, 1.017) | 0.334 |
|  |  |  |  | Penalized weighted | 0.976 (0.958, 0.994) | 0.008 |
| Thyroid | 527/235 | 0.541 | < 0.001 | Inverse variance weighted | 0.994 (0.986, 1.002) | 0.161 |
|  |  |  |  | MR-Egger | 0.99 (0.98, 0.999) | 0.018 |
|  |  |  |  | Penalized weighted | 0.994 (0.984, 1.004) | 0.234 |
| Lung | 1728/757 | 0.840 | 0.205 | Inverse variance weighted | 1.012 (1.004, 1.021) | 0.005 |
|  |  |  |  | MR-Egger | 1.013 (1.002, 1.024) | 0.012 |
|  |  |  |  | Penalized weighted | 1.014 (1.003, 1.026) | 0.017 |
| Breast | 13903/3978 | 0.240 | 0.306 | Inverse variance weighted | 1.003 (1.0005, 1.005) | 0.017 |
|  |  |  |  | MR-Egger | 1.003 (1.0005, 1.005) | 0.007 |
|  |  |  |  | Penalized weighted | 1.003 (1.0005, 1.005) | 0.018 |
| Pancreas | 471/192 | 0.982 | 0.556 | Inverse variance weighted | 1.028 (1.01, 1.046) | 0.002 |
|  |  |  |  | MR-Egger | 1.031 (1.003, 1.06) | 0.016 |
|  |  |  |  | Penalized weighted | 1.031 (1.005, 1.057) | 0.019 |
| Esophagus or stomach | 929/162 | 0.691 | 0.039 | Inverse variance weighted | 0.999 (0.998, 0.9996) | 0.001 |
|  |  |  |  | MR-Egger | 0.999 (0.998, 0.9997) | 0.002 |
|  |  |  |  | Penalized weighted | 0.999 (0.998, 0.9996) | 0.003 |
| Colon | 2897/896 | 0.043 | 0.142 | Inverse variance weighted | 1 (0.998, 1.002) | 0.909 |
|  |  |  |  | MR-Egger | 0.998 (0.996, 1) | 0.037 |
|  |  |  |  | Penalized weighted | 1 (0.997, 1.003) | 0.897 |
| Rectum | 1808/283 | 0.129 | < 0.001 | Inverse variance weighted | 1.001 (0.994, 1.008) | 0.794 |
|  |  |  |  | MR-Egger | 1.009 (1.001, 1.017) | 0.007 |
|  |  |  |  | Penalized weighted | 0.999 (0.988, 1.009) | 0.786 |
| Prostate | 7441/3351 | 0.034 | 0.039 | Inverse variance weighted | 0.998 (0.996, 1.001) | 0.16 |
|  |  |  |  | MR-Egger | 0.998 (0.996, 1.001) | 0.074 |
|  |  |  |  | Penalized weighted | 0.997 (0.994, 0.999) | 0.01 |
| Bladder | 1550/692 | 0.914 | 0.230 | Inverse variance weighted | 0.991 (0.971, 1.011) | 0.37 |
|  |  |  |  | MR-Egger | 0.979 (0.95, 1.009) | 0.091 |
|  |  |  |  | Penalized weighted | 0.991 (0.953, 1.03) | 0.647 |
| Kidney | 1021/317 | 0.370 | 0.054 | Inverse variance weighted | 0.994 (0.983, 1.005) | 0.255 |
|  |  |  |  | MR-Egger | 0.986 (0.971, 1.001) | 0.033 |
|  |  |  |  | Penalized weighted | 0.978 (0.96, 0.996) | 0.018 |
| Cervix | 5998/565 | 0.893 | 0.119 | Inverse variance weighted | 1.006 (1.001, 1.01) | 0.017 |
|  |  |  |  | MR-Egger | 1.006 (1.001, 1.012) | 0.013 |
|  |  |  |  | Penalized weighted | 1.007 (1.001, 1.013) | 0.015 |
| Ovary | 1006/253 | 0.755 | 0.885 | Inverse variance weighted | 1.013 (0.996, 1.031) | 0.143 |
|  |  |  |  | MR-Egger | 1.022 (0.998, 1.047) | 0.033 |
|  |  |  |  | Penalized weighted | 1.017 (0.995, 1.04) | 0.137 |
| Endometrium | 1414/623 | 0.814 | 0.722 | Inverse variance weighted | 1.016 (1.009, 1.023) | < 0.001 |
|  |  |  |  | MR-Egger | 1.013 (1.001, 1.025) | 0.009 |
|  |  |  |  | Penalized weighted | 1.015 (1.005, 1.025) | 0.004 |
| Lymphocytic leukemia | 594/258 | 0.672 | 0.244 | Inverse variance weighted | 0.984 (0.967, 1.002) | 0.081 |
|  |  |  |  | MR-Egger | 0.975 (0.948, 1.002) | 0.038 |
|  |  |  |  | Penalized weighted | 0.963 (0.936, 0.99) | 0.007 |
| Melanoma | 4271/2506 | 0.883 | 0.830 | Inverse variance weighted | 1.021 (1.01, 1.032) | < 0.001 |
|  |  |  |  | MR-Egger | 1.02 (1.003, 1.036) | 0.009 |
|  |  |  |  | Penalized weighted | 1.024 (1.008, 1.04) | 0.004 |
| Non-Hodgkin lymphoma | 1760/640 | 0.610 | 0.862 | Inverse variance weighted | 0.995 (0.992, 0.999) | 0.005 |
|  |  |  |  | MR-Egger | 0.995 (0.991, 0.999) | 0.005 |
|  |  |  |  | Penalized weighted | 0.994 (0.99, 0.998) | 0.007 |

OR = odds ratio, CI = confidence interval, GERA = Genetic Epidemiology Research on Adult Health and Aging
The effect sizes of the causal estimates were from one standard deviation increase in the genetic predisposition for atrial fibrillation.

**Figure 1.**
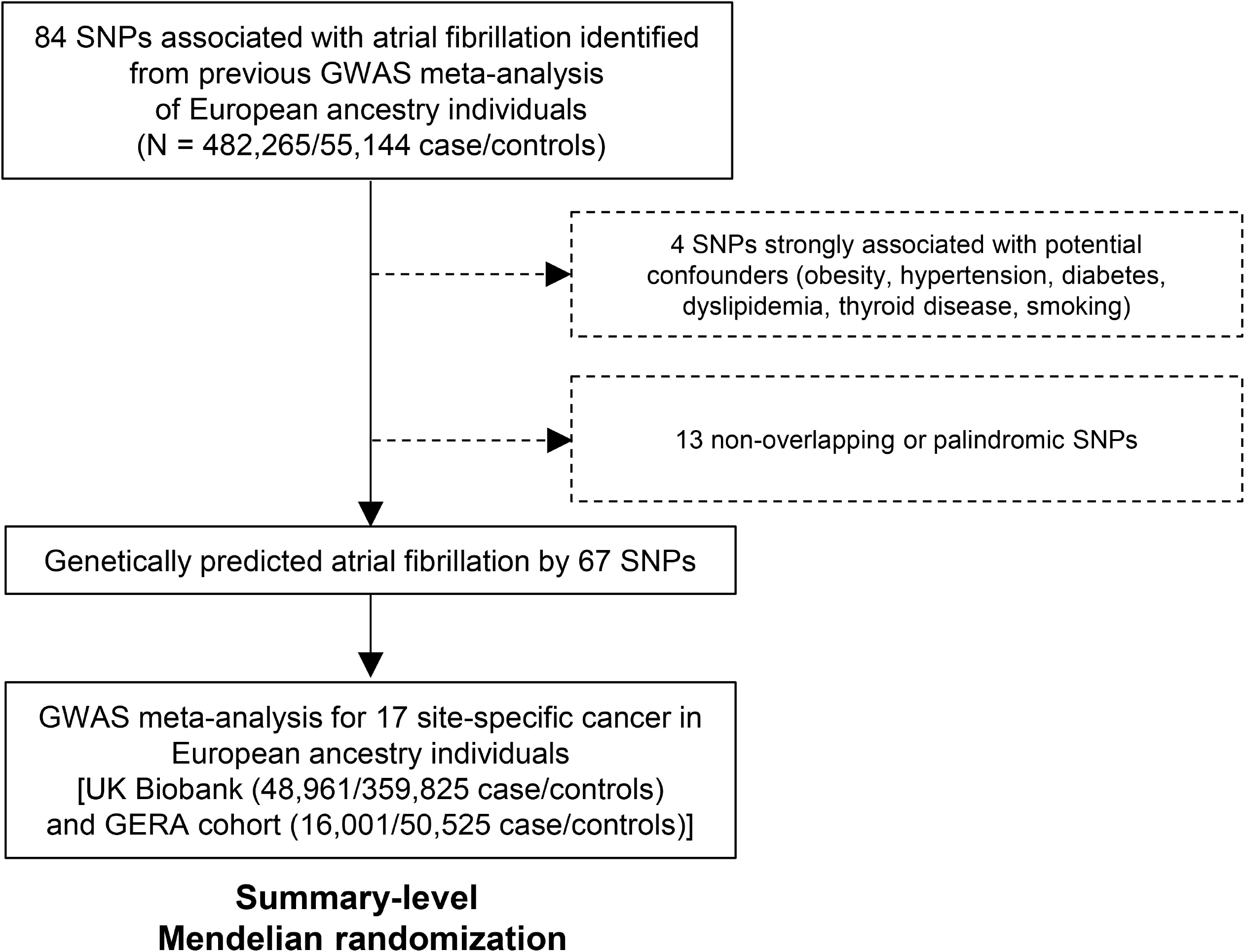
Study flow diagram. SNPs = single nucleotide polymorphism, GWAS = genome-wide association study, GERA = Genetic Epidemiology Research on Adult Health and Aging.

**Figure 2.**
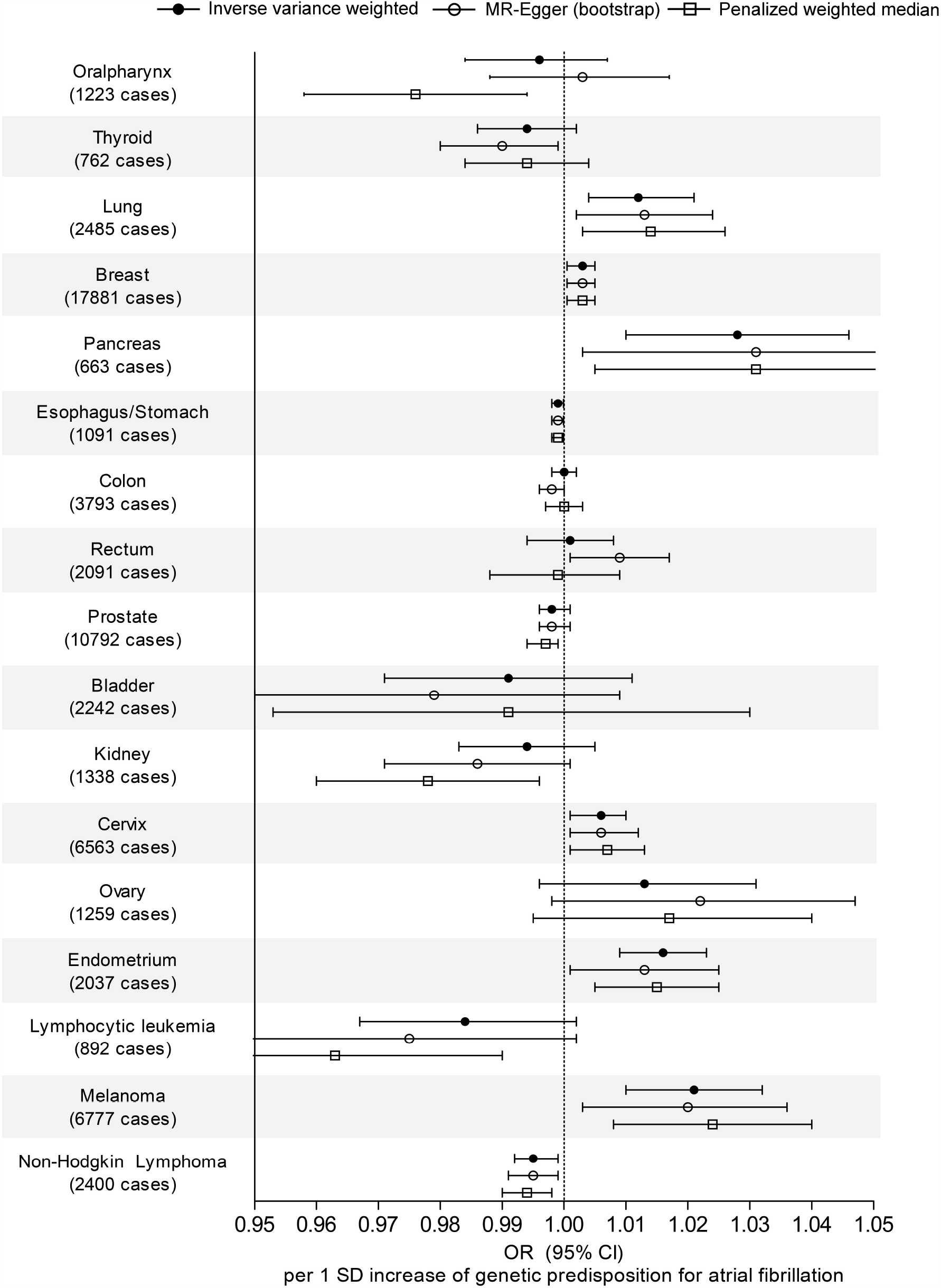
Causal estimates from kidney function on site-specific cancer risk. The x-axes indicate the odds ratios and confidence intervals, from one standard deviation increase in the genetic predisposition for atrial fibrillation. The vertical broken line indicates 1 OR. The black round causal estimates are from the multiplicative random effect inverse variance weighted method. The white round causal estimates are from the MR-Egger regression with bootstrapped standard error. The white square causal esimtates are from the penalized weighted median method.

The causal estimates for pancreas cancer risk indicated that genetical predisposition for AF may be significantly associated with higher risk of pancreas cancer. However, the findings were non-significant in the sensitivity analysis reducing the number of SNPs by applying more stringent threshold.

On the other hand, higher genetic predisposition for AF was significantly associated with lower risk of esophagus/gastric cancer. The results were consistent throughout the performed sensitivity analyses. However, MR-Egger test for directional pleiotropy indicated that significant pleiotropic effect was detected in the causal estimate for the risk of esophagus or stomach cancer.

The genetically predicted AF was significantly associated with non-Hodgkin lymphoma in the main analysis, but the significance was attenuated when we included fewer but stronger SNPs as the genetic instrument considering the overlapping sample issue.

## Discussion

In this MR analysis, we identified that AF may cause cancers of lung, breast, cervix, endometrium, and melanoma. The genetic predisposition of AF was significantly associated with lower risk of esophagus/gastric cancer, but possibility of a directional pleiotropy remained in the association. The findings suggest that AF is a causative factor for certain cite-specific cancer risks.

The previous observational findings suggested bidirectional association between AF and cancer. The studies reporting that AF was higher incident cancer risks raised question whether AF may have causal effect on certain risks of malignancies.^5,25^ However, as delayed diagnosis for AF and cancer is possible and the diseases share common risk factors, the presence of causal effect of AF on cancer was only suspected but yet confirmed.^26^ In this MR analysis, we constructed genetic instrument for AF by implementing the largest GWAS result to date and applied to one of the largest GWAS meta-analyses for site-specific cancer risks by the summary-level MR. With our efforts to attain the key assumptions of MR, we identified that AF causally affects risks of certain site-specific cancers, which has been newly identified by this methodology.

Our findings may serve as an evidence for cancer screening guideline for AF patients. In AF patients, additional screening may be necessary for lung, breast, cervix, endometrium, and melanoma cancers because AF causally elevates the cancer risks. Or, screening of AF may be considered in the individuals with such cancers, as subclinical AF is common and may affect cancer progression considering its’ present effect on cancer development. As non-cardiovascular mortality in AF patients is common and largely attributed by malignancies,^27^ such strategy may ameliorate the burden of cancer-related complications and risk of mortality in AF patients by promoting early diagnosis.^26,28^

Our study encourages future studies for the mechanism regarding the identified causal effects. It is well described that AF causes systemic hypercoagulability and directly, formation of atrial thrombosis causing arterial thromboembolism.^29,30^ As patients with primary venous thromboembolism showed higher risk of cancers without evidence of occult malignancy, vascular thrombosis has been suspected to have causal effect on cancer development.^31,32^ In addition, two studies reported that not only venous, but also arterial thrombosis was associated with increased risk of incident cancer identification, from 746862 cancer patients in the United States^33^ and from 6600 Danish patients diagnosed for low limb arterial thrombosis.^34^ Although the possibility of delayed-diagnosis for occult malignancy after identification of arterial thrombosis cannot be disregarded, certain portion of the incident cancer events might have been related to causal effects from impaired circulation due to vascular thrombosis. Such hypothesis was supported by the previous observational findings suggesting anti-tumor effect of anti-thrombotic drugs.^26,28,35^ Linkage between hypercoagulability state and cancer risk has been recently suggested.^36^ Therefore, that AF provoking vascular thrombosis, hypercoagulability, and impaired circulation may be the vertical pathway of the causal effect from AF on risk of cancer development. A future study targeting the thrombogenic pathway as an etiology of cancer development is warranted. In addition, the mechanism that the causal effects were apparent for specific cancer types should be studied. Furthermore, if the biologic pathway is relevant, additional trial may test the efficacy of appropriate usage of preventive anticoagulative therapy or prompt rhythm control for reducing the cancer risks in AF patients.^28,37^ On the other hand, AF may have causal effect on risk of certain types of cancer through undetermined pathway, and hemodynamic effect or neurohormonal response related to AF may be investigated as a potential mechanism.

That genetic predisposition for AF was associated with lower risk of gastric/esophageal cancer needs to be explained, although the identification for direct mechanism is beyond the current study’s possibility. As AF directly reducing a cancer risk is biologically less plausible, it possible that, as cancer cases are determined by the first cases in the previous study, an early diagnosis of AF-affected cancers might have led to under inclusion of gastric-esophageal cancer cases in the outcome data. Or, considering the significant MR-Egger intercept P value indicating presence of a directional pleiotropy, group of the instrumented genetic variants might have acted through a pleiotropic pathway, which is a violation of the InSIDE assumption and causes false-positive findings even in the pleiotropy-robust sensitivity analyses. A future study is warranted to confirm the findings with additional dissection of the cancer subtypes, as various cancer types with different histology may occur in esophagus and stomach.

There are several limitations in this study. First, this study does not confirm whether appropriate management for AF, by rhythm control or by preventive antithrombotic treatment, may reduce cancer risks. Additional trial targeting the identified causal pathway is necessary to extend the clinical application of our findings, along with a study investigating the mechanistical explanation for the results. Second, the reason for the causal effects are different according to cancer types was not explained in this study. As a false-negative bias is possible in MR, as genetic instrument explains a portion of the exposure of interest, other cancer risks may also be affected from AF. Yet, the identified cancers that are causally affected from AF may be prioritized for future studies investigating the efficacy of management of AF on reducing risk of a type of cancer. Third, the clinical effect size of AF on cancer risks may be different from the identified genetic effects which were small.^38^ The genetic effect sizes from MR might have been shrunken due to the availability of genetic information to explain AF and the relatively small cases of each cancer compared to the controls. Or it is possible that AF may actually have relatively small effect on cancer risk than that from other common etiologies of malignancy. Fourth, due to the limitation of the available genetic information, detailed subtypes of atrial fibrillation (persistent or paroxysmal) or cancer (histologic classification) were not considered. Lastly, the study population was limited to European ancestry individuals, thus, generalizability of our finding is not secured for other ethnic populations. In conclusion, AF is a causal factor for certain types of cancer. Appropriate cancer screening should be suggested in clinical guidelines for AF patients. Future trial is necessary to confirm whether appropriate management of AF may reduce the risk of cancer which is a major cause of deaths in AF patients.

## Data Availability

The main data for this study are archived in public repositories by previous studies.

## Acknowledgements

The study was based on the data provided by the UK Biobank consortium (application No. 53799). We thank the investigators of the previous studies who provided the valuable genetic summary statistics for this study.

## Data Availability Statement

The main data for this study are archived in public repositories by previous studies.

