## Supplemental Table 1 for "Reverse causal effect of atrial fibrillation on 17 site-specific cancer risk: A Mendelian randomization study"

Sehoon Park et al.

**Supplemental Table 2.** Sensitivity analysis including 47 SNPs with stronger association (P < 1×10^-11^) with phenotypical atrial fibrillation as the genetic instrument in concerns of overlapping sample issue for the significant findings in the main analysis.

| Site-specific cancer | Cochran’s Q statistics P value | MR-Egger test P value for directional pleiotropy | MR method | OR (95% CI) | P |
| --- | --- | --- | --- | --- | --- |
| Lung | 0.892 | 0.069 | Inverse variance weighted | 1.013 (1.004, 1.022) | 0.004 |
|  |  |  | MR-Egger | 1.012 (1, 1.024) | 0.029 |
|  |  |  | Penalized weighted median | 1.013 (1.002, 1.025) | 0.024 |
| Breast | 0.436 | 0.290 | Inverse variance weighted | 1.003 (1.001, 1.005) | 0.012 |
|  |  |  | MR-Egger | 1.003 (1.001, 1.005) | 0.003 |
|  |  |  | Penalized weighted median | 1.003 (1.0004, 1.005) | 0.021 |
| Pancreas | 0.806 | 0.136 | Inverse variance weighted | 1.022 (0.993, 1.053) | 0.142 |
|  |  |  | MR-Egger | 1.033 (0.996, 1.071) | 0.037 |
|  |  |  | Penalized weighted median | 1.023 (0.987, 1.059) | 0.215 |
| Cervix | 0.971 | 0.235 | Inverse variance weighted | 1.008 (1.003, 1.012) | 0.001 |
|  |  |  | MR-Egger | 1.007 (1.002, 1.013) | 0.005 |
|  |  |  | Penalized weighted median | 1.007 (1.001, 1.013) | 0.016 |
| Endometrium | 0.824 | 0.952 | Inverse variance weighted | 1.017 (1.007, 1.027) | 0.001 |
|  |  |  | MR-Egger | 1.017 (1.004, 1.031) | 0.003 |
|  |  |  | Penalized weighted median | 1.017 (1.004, 1.029) | 0.009 |
| Melanoma | 0.881 | 0.645 | Inverse variance weighted | 1.020 (1.009, 1.032) | < 0.001 |
|  |  |  | MR-Egger | 1.019 (1.001, 1.038) | 0.008 |
|  |  |  | Penalized weighted median | 1.022 (1.005, 1.039) | 0.011 |
| Esophagus  or stomach | 0.473 | 0.126 | Inverse variance weighted | 0.999 (0.998, 0.9997) | 0.003 |
|  |  |  | MR-Egger | 0.999 (0.998, 0.9997) | 0.001 |
|  |  |  | Penalized weighted median | 0.999 (0.998, 0.9998) | 0.008 |
| Non-Hodgkin lymphoma | 0.583 | 0.062 | Inverse variance weighted | 1.001 (0.991, 1.012) | 0.790 |
|  |  |  | MR-Egger | 0.927 (0.856, 1.004) | 0.069 |
|  |  |  | Penazlied weighted median | 1.001 (0.985, 1.016) | 0.943 |

OR = odds ratio, CI = confidence interval.

The effect sizes of the causal estimates were from one standard deviation increase in the genetic predisposition for atrial fibrillation.
